# Impact of Primary Care Team Configuration on Access and Quality of Care

**DOI:** 10.1101/2023.05.18.23290117

**Authors:** Sylvia J. Hysong, Kelley Arredondo, Houston F. Lester, Richard SoRelle, Trang Pham, Frederick L. Oswald, LeChauncy Woodard, Laura A. Petersen, Joshua Hamer, Ashley M. Hughes

**Author notes:** Corresponding author (SH).

## Abstract

**Importance:** The *Joint Principles of the Patient Centered Medical Home* (PCMH) call for a team-based approach to delivering primary care – however, they provide little guidance on what should be the optimal staffing configuration to best achieve care objectives. Given recent primary care physician reports of higher intentions to leave primary care because of workload concerns, configuring primary care correctly to deliver high-quality, accessible care equitably without losing clinicians along the way is paramount.

**Objective:** This paper aims to empirically examine the extent to which variations in team configurations within PACTs predict primary care access and quality.

**Design:** Prospective, observational database review of Veterans Health Administration (VHA) Corporate Data Warehouse measures describing staffing configuration and clinical performance (access, quality) of primary care teams. We extracted monthly data from February and December 2020.

**Setting:** VHA medical centers and community-based outpatient clinics nationwide.

**Participants:** 22,392 primary-care personnel representing 7,750 PACTs from 1,050 VHA healthcare facilities nationwide.

**Exposure:** Adherence to a VHA-recommended primary care team configuration of one primary care provider, registered nurse, licensed vocational nurse, and administrative clerk, respectively. Using network analysis methods we calculated, for each team, an overall adherence score and two team network characteristics (degree, Blau’s index) to capture role diversity and clinician assignment to multiple teams. We also calculated team size and number of full-time equivalents (FTE).

**Main Outcome and Measures:** Access to care and quality of care, as measured by the following outcomes: 1) Average third next available appointment (2) ER/urgent care (UC) utilization rate (3) Inbound to total outbound primary care secure messages ratio); (4) Team 2-day post-discharge contact (5) Hemoglobin A1c control (an indicator of poor diabetes management) (6) Diabetic nephropathy screening and (7) Hypertension control).

**Results:** Adherence to the recommended configuration as measured by the adherence index, had different outcomes, both pre- and post-onset of the COVID pandemic. Pre-pandemic onset, overall adherence significantly predicted no outcomes. However, individual network characteristic analysis showed increased role diversity was associated with decreased ER/UC utilization and greater patient engagement through secure messaging. Larger teams exhibited improved 2-day post-hospital discharge contact, but worse access in terms of third next available appointments.

Post-pandemic onset, teams with lower overall adherence showed higher ER/UC utilization. Higher multiple-team membership was associated with lower ER/UC utilization. Larger teams exhibited lower ER/UC utilization scores, but lower 2-day post-discharge contact and nephropathy screening scores. In nearly all cases, however, teams with larger numbers of FTEs were associated with better outcomes.,

**Conclusions and Relevance:** Primary care teams require a minimum amount of FTE capacity to deliver high quality and access to health care. Future work should examine the impact of staffing levels by specific job role to further optimize staffing configurations.

## Background

The *Joint Principles of the Patient Centered Medical Home* (PCMH) call for a team-based approach to delivering first contact (i.e., primary care), continuous, and comprehensive care. Within the *Principles*, a consistent team of healthcare professionals led by a provider *collectively* take responsibility for the ongoing care of patients. Through this approach patients are purported to receive well-coordinated, high-quality, whole-person care.^1^ The *Principles* are silent, however, on what should be the configuration of staffing to best achieve these care objectives.

The Veterans Health Administration (VHA)’s adaptation of the PCMH principles (coined Patient Aligned Care Teams or PACTs) transformed primary care services throughout clinics nationwide. As part of PACT adoption, the VHA recommended staffing PACTs with a core ‘teamlet’ consisting of one registered nurse care manager, one licensed vocational nurse, and one scheduling clerk per primary care provider, each at full-time effort. This recommended staffing configuration-- a team consisting of four healthcare professionals with distinct yet complementary skills, professional backgrounds, and responsibilities – made conceptual sense and was thought to situate primary care teams well to meet their stated care objectives. ^2^

Early research on adoption of PCMH principles through PACT-based care indicated multiple benefits, including higher patient satisfaction and clinical quality, as well as lower staff burnout, hospitalization rates for ambulatory care-sensitive conditions, and emergency room (ER) utilization, respectively.^3^ Further research suggested some PACT benefits may operate through the diversity of staff roles included in the teams (i.e., distribution of physicians, nurses, clerks, and other professionals), as more role diversity has been linked to increased team performance.^4^

Real-world implementation of the PACT model, however, varies widely and significantly in team structure and staffing. This in turn may introduce unwanted variability in quality of care and downstream effects on patient outcomes. For example, a 2014 analysis of PACT staff membership indicates (1) only 19% of primary care PACTs adhered to VA’s recommended configuration (described above) and (2) configurations of the remaining 81% of PACTs varied widely.^4^ More recent empirical work in this area shows PACTs with team members who are shared across other PACTs (i.e., multi-team membership) score worse on access to primary care metrics (e.g., increased patient ER utilization).^5^ This is just one way in which PACTs can deviate from the recommended staffing structure since the recommendation is that each team-member be assigned to one team at full-time effort.^5^ Given how recent primary care physician surveys report higher stress, burnout, and intentions to leave primary care because of workload concerns,^6^ configuring primary care correctly to deliver high-quality, accessible care equitably amongst communities of all levels of need without losing clinicians along the way is paramount. Nevertheless, we are unaware of studies examining how variations in PACTs may directly impact quality of the care delivered.^7^ Therefore, this paper aims to empirically examine the extent to which variations in team configurations within PACTs predict primary care access and quality. Specifically, we hypothesize that PACTs adhering more closely to the recommended team configuration described above will exhibit higher primary care access and clinical quality than those more strongly deviating from the recommended team configuration.

## Methods

### Design, Setting, and Sample

As part of a larger, longitudinal database review,^8^ we conducted a prospective observational study and applied methods from network analysis (NA) to answer our research question. We used clinical and administrative VHA data sources (described below) to examine the access to and quality of primary care delivered by PACTs at VA Medical Centers (VAMCs) and Community-Based Outpatient Clinics (CBOCs) nationwide. Due to the diversity of team configuration implementation across VHA facilities nationwide, many types of team configurations can be examined efficiently and effectively, thus creating a natural laboratory for studying staffing.^9-11^ We examined the records of 22,392 primary-care personnel representing 7,750 PACTs from 1,050 VHA healthcare facilities nationwide (VAMCs and CBOCs). Teams that did not directly focus on primary care (e.g., mental health teams), and teams created for educational purposes (e.g., resident-led teams) were excluded.

## Data Sources

### Primary Care Almanac Team Assignments Report

The Primary Care Almanac Team Assignments Report (TAR) displays all active PACTs at every VAMC and CBOC within the VHA system, along with the names and roles of the primary care staff members assigned to each team. It is updated nightly and created from fields within the Corporate Data Warehouse (CDW), thereby facilitating linkage to other data sources for our study. We used the TAR to identify each team and calculate its respective network properties.

### Data Sources for Clinical Performance

Clinical performance was calculated from multiple sources, described below.

#### Corporate Data Warehouse

The VHA CDW is a national repository of clinical and administrative systems; it is a relational database organized into a collection of clinical and administrative data domains, ranging from October 1999 to present.

#### EQM Measures

Clinical performance was measured monthly using indicators from outpatient electronic quality measures (eQM) from the VHA’s Office of Analytics and Performance Integration. This subset of clinical quality measures relies on nationwide, automated extraction of data pooled in the VHA CDW to generate near real-time, full-population measures of clinical performance, which are updated daily.^12^ Scores are calculated using the entire patient population (i.e., 100% sampling) rather than estimating scores based on a sample of abstracted records to reduce sampling and missing-data concerns.

#### HEDIS Metrics

HEDIS is a tool used by more than 90 percent of America’s health plans to measure performance on important dimensions of care and service. Altogether, HEDIS consists of 81 measures across 5 domains of care, including effectiveness of care (i.e., clinical performance). HEDIS Metrics for VHA facilities are available through the Strategic Analytics for Improvement and Learning reporting system within the VHA.

## Measures

### Adherence to Recommended Team Configuration

Network analysis, a methodology that describes and analyzes the configural and dynamic properties of a network, has become a popular and useful tool for empirically relating team configurations to health care outcomes. ^4,5,13,14^ We employed NA to determine the extent to which actual and ideal team membership configurations are aligned (described below). Following Crawford and colleagues’ methods,^5^ we examined two team network characteristics: team-level *degree* (i.e., the average number of teams to which each member of a given team is assigned) and *Blau’s index*^14^ (i.e., the extent to which the team contains diversity of roles [primary care provider, registered nurse care manager, licensed vocational nurse, and scheduling clerk]). A Blau’s index of zero indicates complete homogeneity; complete heterogeneity is a function of the number of available categories (in this case roles) and the distribution of members in each category. In the case of PACTs, a team that follows VHA’s recommendation of exactly one provider, one registered nurse care manager, one licensed vocational nurse, and one scheduling clerk where each team member is assigned the same team and only that team mathematically has a degree of 1 and a Blau’s index of .75. Both must be true to satisfy the VHA’s recommended PACT configuration.

Additionally, we examined *team size* (i.e., the number of members on a team) and *team full-time equivalence* (FTE, i.e., the sum of the number of team members multiplied by their respective work effort, expressed as a percent of a 40-hour work week). A recommended PACT configuration contains an FTE of 4: four individuals, each working full time (see Team 6 in Figure 1 for an illustrative example).

**Figure 1.**
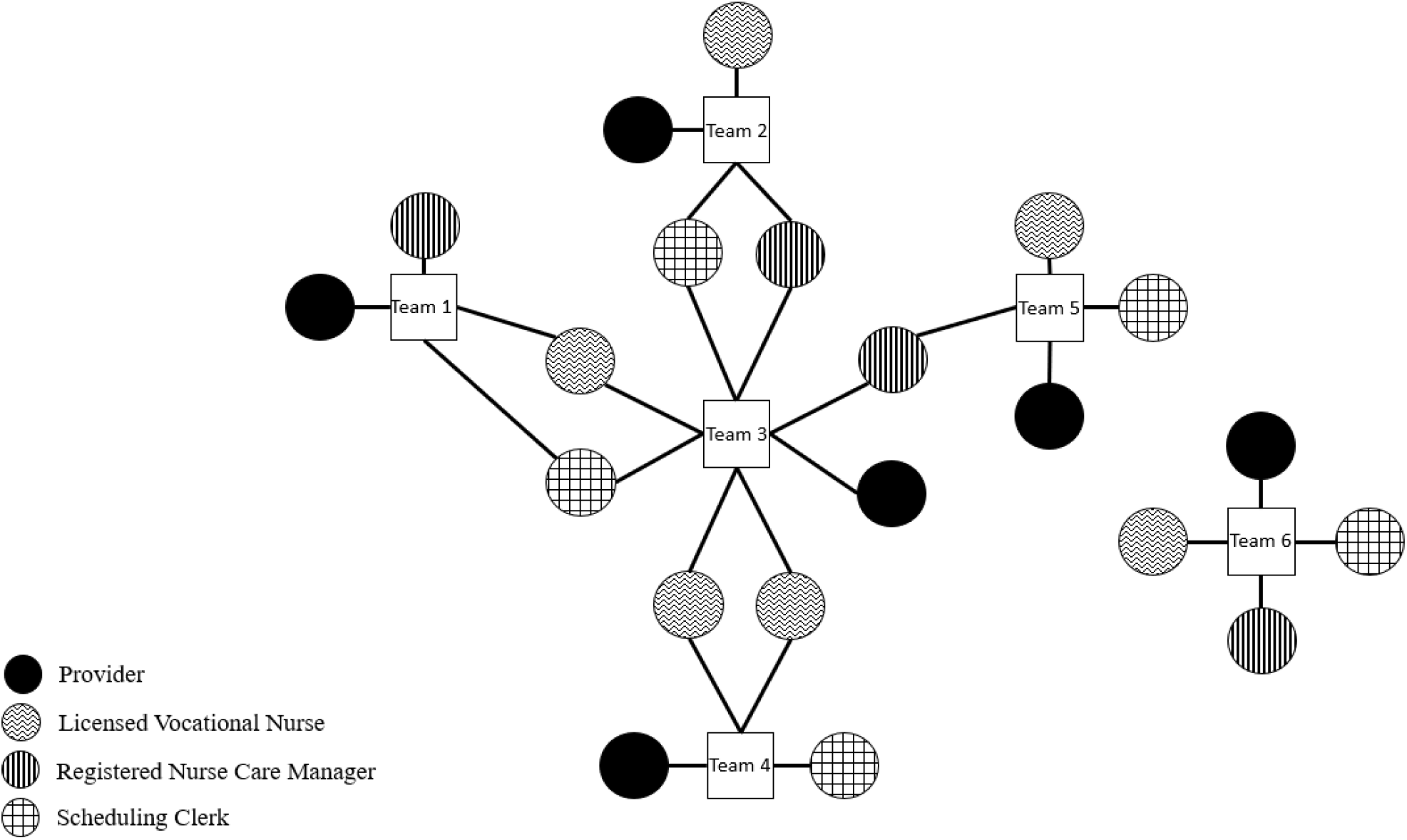
Network diagram of sample primary care team staffing configurations. Note. Nodes (circles and squares) depict primary care teams (white squares) and their respective members (circles). Edges (lines) connect members to their respective team(s); circles with more than one edge represent clinicians assigned to more than one team. Each circle’s texture pattern indicates a different team role.

To quantify overall adherence to the recommended team configuration we calculated an *adherence index* using the afore-described NA configural properties. For each team, we calculated degree and Blau’s index, subtracted each from its respective recommended score, and used this as a team deviation score for each property. We then calculated the average of these deviations to obtain an overall measure of adherence. Figure 1 depicts various existing VHA PACT configurations. The figure illustrates the recommended configuration (depicted in Team 6) and how PACTs can deviate from the recommended configuration by having team-members who are part of multiple teams (e.g., all members of Team 3, except the provider), and by having lower role diversity (e.g., Team 4).

### Clinical Performance

The specific clinical performance outcomes examined, which include measures of access to care and quality of care, were selected by an expert panel focusing on metrics that are of the highest value to primary care and available at the team level. Details on the selection process (and resulting metrics) are published elsewhere.^15^ Based on the expert panel findings, we extracted from the aforementioned data sources the following access-to-care outcomes: (1) Average third next available appointment (2) ER/urgent care (UC) utilization rate (3) Inbound to total outbound primary care secure messages ratio); and the following care quality outcomes: (4) Team 2-day post-discharge contact (5) Hemoglobin A1c control (an indicator of poor diabetes management) (6) Diabetic nephropathy screening and (7) Hypertension control).

### Procedure

We extracted monthly data from February 2020 (before the World Health Organization declared COVID-19 as a pandemic) and December 2020 (after declaring COVID-19 a pandemic, but before wide vaccine availability). Though not our primary phenomenon of interest, observing these two periods was important because the working conditions for primary care changed materially during the pandemic, potentially leading to differential effects of team configuration under ordinary vs. extreme (pandemic) circumstances. We specifically selected December 2020 as the COVID period of interest because this reflected a point in time when (a) COVID would have been widespread in the US;^16^ (b) primary care would have had time to adjust to COVID-related changes; and (c) would have still operated under significant restrictions due to the absence of a widely available vaccine.^17^

Datasets were imported into a SQL-friendly relational database platform, so they could be linked to one another using TAR identifiers. Data were aggregated to the team level. All team configuration measures were calculated as described above from fields in TAR.

### Analysis

We conducted two sets of analyses to test our hypotheses: First, we conducted a series of multi-level models (MLM) with each measure of team-level clinical performance as the outcome (each outcome modeled separately), and the following variables as team-level (level-1) predictors: overall deviation from ideal team configuration (adherence index), total team FTE, team size; facility was modeled as a random effect (level-2) to account for facility clustering (i.e., to determine whether the teams differ across facilities). To examine the relative contributions of individual configural PACT characteristics more closely, we then conducted a second series of MLM with each NA component (degree and Blau’s index) modeled separately: each measure of clinical performance (modeled separately) served as the outcome; team degree, team Blau’s index, team size, and team FTE served as team-level (level-1) predictors. A random intercept due to facility was also included to account for between-facility mean differences in the outcomes of interest.

## Results

Table 1 summarizes characteristics of the teams and primary care personnel involved.

**Table 1.**
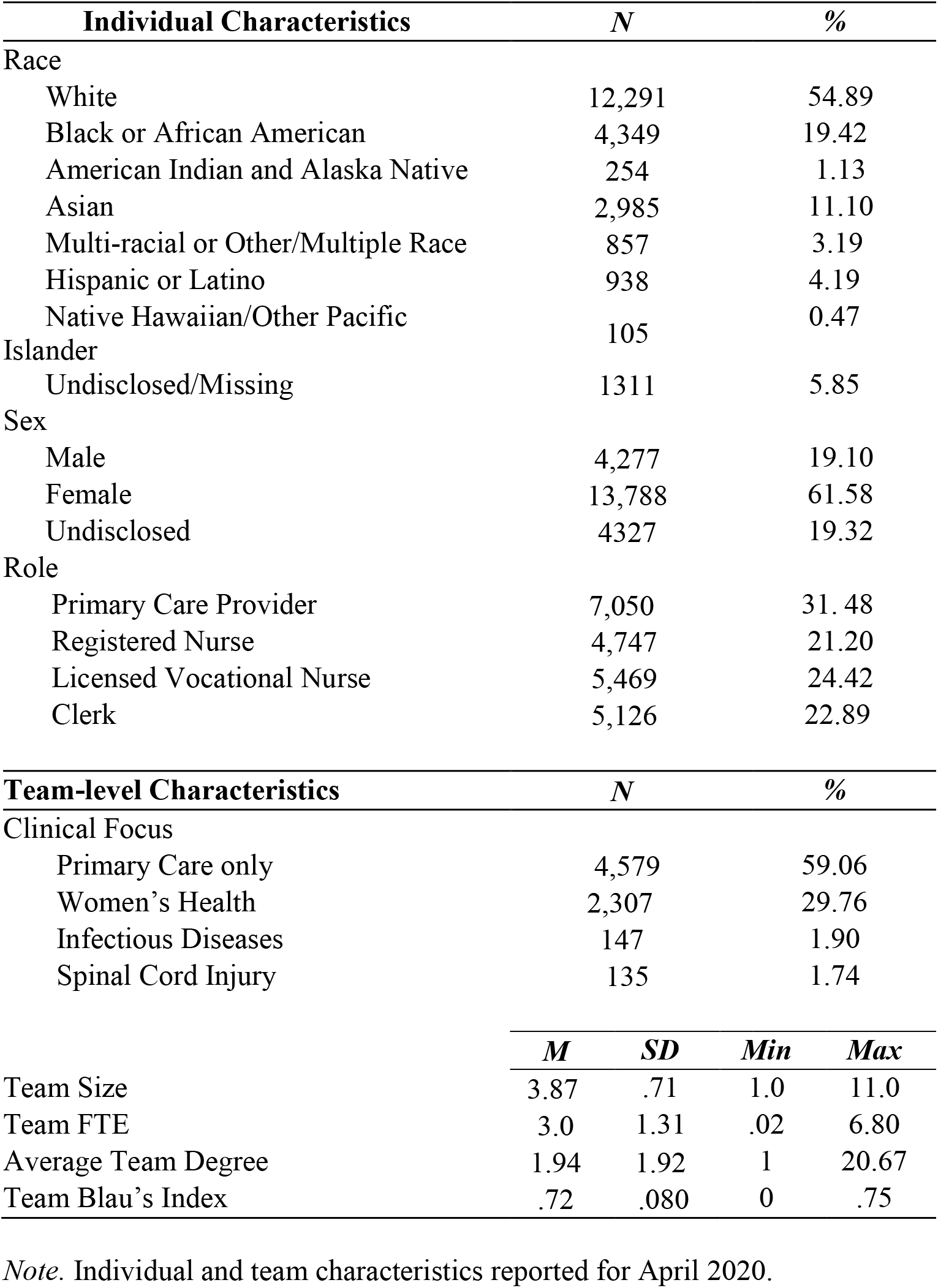
Characteristics of VHA primary care team members and PACTs

Adherence to the recommended configuration as measured by the adherence index, had different outcomes, both pre- and post-onset of the COVID pandemic. Pre-pandemic onset, adherence to the recommended configuration was not significantly associated with any outcomes. Tests of individual network characteristics, however, painted a more complex picture: greater deviation from the expected Blau index of .75 (i.e., less role diversity) was associated with decreased ER/UC utilization, suggesting better access for patients cared for by PACTs with less role diversity than for patients assigned to more role-diverse PACTs. However, teams with lower role diversity showed worse patient engagement through secure messaging communication ratios (i.e., patients were less likely to get a response from clinicians in less role-diverse PACTs). Greater team size (i.e., teams with more members) was associated with improved 2-day post-hospital discharge contact scores (a measure of care coordination with primary care) but worse access in terms of third next available appointments. Of note, 21% of teams had greater than 4 members, the recommended size, indicating most deviations from the recommended team size of 4 were due to understaffing.

Post-pandemic onset, PACTs with less overall adherence to the recommended configuration showed higher ER/UC utilization (lower is better). Closer examination of individual configural characteristics showed PACTs with a higher degree (i.e., multi-team membership (MTM)) were more likely to exhibit higher ER/UC utilization amongst their patients, compared to PACTs with lower degree. Further, greater deviation from the expected Blau index (i.e. less role diversity) was associated with less ER/UC utilization. Greater team size was associated with less ER/UC utilization; however, greater team size was also associated with lower care quality (i.e., team 2-day post-discharge contact and nephropathy screening scores).

Table 2 provides a complete summary of these findings.

**Table 2.** Summary of parameter estimates (B) and standard errors resulting from multi-level model analyses.

| Outcome |  | Blau |  | Degree |  | Team Size |  | Overall Deviation |  | FTE |  | Intercept |  |
| --- | --- | --- | --- | --- | --- | --- | --- | --- | --- | --- | --- | --- | --- |
| *Pre vs Post-COVID onset* |  | *Pre* | *Post* | *Pre* | *Post* | *Pre* | *Post* | *Pre* | *Post* | *Pre* | *Post* | *Pre* | *Post* |
| 3rd Next Available | *B* | 5.21 | .62 | -.05 | .44 | **2.30** | .25 | <.01 | .44 | **-1.28** | **-1.00** | 21.42 | 14.84 |
|  | *SE* | 1.01 | 5.22 | .58 | .31 | **1.19** | .77 | .57 | .31 | **.68** | **.41** |  |  |
| ER/UC Utilization | *B* | **-35.56** | **-16.60** | 1.11 | **2.15** | -1.72 | **-1.39** | .65 | **1.57** | **-3.12** | **-1.02** | 34.85 | 26.78 |
|  | *SE* | **10.85** | **7.32** | .74 | **.65** | 1.45 | **1.02** | .37 | **.60** | **.54** | **.47** |  |  |
| Team 2-day ratio | *B* | .25 | -.13 | -.01 | -.01 | **.04** | **.05** | <-.01 | <-.01 | **.02** | <.01 | .57 | .52 |
|  | *SE* | .22 | .18 | .01 | .01 | **.02** | **.02** | .01 | .01 | **.01** | .01 |  |  |
| UC Utilization | *B* | -.02 | -.02 | -.01 | <-.01 | <.01 | <.01 | <.01 | <.01 | **-.04** | **-.004** | .05 | .04 |
|  | *SE* | .02 | .02 | .001 | .002 | .003 | .002 | .00 | .00 | **.001** | **.001** |  |  |
| Ratio of in/out messages | *B* | **6.02** | 1.81 | -.03 | .17 | -.06 | **-.44** | .08 | .21 | .12 | .17 | 1.63 | 1.48 |
|  | *SE* | **1.83** | 1.90 | .12 | .13 | .18 | **.20** | .12 | .13 | .10 | .12 |  |  |

*Note: Numbers in parentheses indicate standard errors. Numbers in **bold type** indicate statistically significant differences in the clinically desired direction (e.g., lower ER/UC utilization is better). Numbers in **bold italics** indicate statistically significant differences in the clinically undesirable direction (e.g., higher ER/UC utilization is worse).*

## Discussion

This study investigated the effectiveness of PCMH-derived team staffing configurations at improving access to and quality of primary care. The study adds to the body of knowledge on PCMH effectiveness by testing the impact of adherence to PCMH staffing principles on metrics of care quality derived from a national expert panel, and by using a national sample of VHA primary care teams, one of the largest available samples in healthcare team research. We found little evidence to support the specific recommended configuration of one primary care provider, one registered nurse care manager, one licensed vocational nurse, and one scheduling clerk under ordinary conditions. However, stricter adherence to the recommended PACT configuration after the onset of COVID was associated with reduced ER/UC utilization, which was both an indicator of patient access to care and a critical resource in the height of the pandemic. Although the overall PACT configuration was not generally associated with the access to and care quality outcomes, analyzing specific PACT staffing components gave further insight into the mechanisms that lead PACT configurations to impact access to and quality care.

In examining specific components of PACT staffing adherence, we found mixed association between configural characteristics of primary care teams and our observed outcomes. However, we did identify some components that had a significant impact on access to and care quality. For example, access to care was more significantly impacted — albeit with mixed results —in teams with more members, regardless of configuration. Larger teams may have achieved better outcomes through having sufficient personnel rather than a strict adherence to a particular team size or staffing mix. One consistent driver of both access and quality, however, was FTE: For all significant relationships, teams with greater FTE exhibited better outcomes, both pre- and post-pandemic onset. Given the mean FTE was 3.02, and only 20% of teams had more than 4 team members (with a maximum FTE of 6.6), our observed findings illustrate the detrimental effects of understaffing in primary care, consistent with other health services research.^6^

Furthermore, our findings suggest that different features of the recommended configuration such as size, role diversity (Blau index), team cohesion (degree), and staffing availability (degree, FTE) differentially impact different aspects of care, especially pre-COVID. For example, the size of the team and FTE of staffing assigned to the PACT appeared to influence care coordination (2-day post-discharge contact ratio) such that smaller teams and teams with greater FTE performed better on care coordination as opposed to larger teams and teams with less FTE per person on a team. One reason for this difference may have to do with MTM, where personnel are assigned to multiple teams for low FTE, which may complicate coordination efforts^18^ and has been associated with lower team performance in a pre-COVID study.^5^ In contrast, team size (i.e., number of individuals in the team) was the only configural feature that did not significantly impact ER/UC utilization. If that is true, then adherence to the recommended configuration may help teams deliver better access by the subsets of benefits each feature yields, rather than by the Gestalt of a singular configuration. This also may explain why, when examined in aggregate, overall adherence to/deviation from the recommended configuration appears to be mostly unrelated to outcomes. Findings also highlight the importance of implementing broader policies to mitigate the current nurse and physician shortages, exacerbating the understaffing effects observed here, especially in rural and health profession shortage areas.^19,20^

## Limitations

Our study is limited by the availability of existing data in our data sources. Certain aspects of quality, such as 30-day readmissions and ambulatory care sensitive conditions, are not available at the team level and could not be assessed for this analysis, despite being common measures of quality in primary care. Similarly, other team-level care quality metrics identified by experts were recommended for adoption (e.g., UC utilization rate adjusted for clinical reason, rates of inappropriate prescribing, number of missed opportunities for care coordination, and timely communication of clinical information within clinic) that do not yet exist as trackable metrics in current systems. As such, the current study may present an incomplete picture of primary care access and quality. However, by including 7,750 teams nationwide, data used in this study represents one of the largest known samples of primary care teams, as the average sample size for studies of teams is 114 teams.^21^ Therefore, our findings, which did not sample all dimensions of access and care quality, nevertheless have ample power for reliable results in the areas sampled.

## Conclusion

Primary care teams require a minimum amount of FTE capacity to deliver the level of access to health care and quality expected of them without danger of clinician burnout and turnover.^19,20^ In addition, staffing primary care teams with specific configural features may yield improvements in different aspects of access and quality. It is therefore important to staff primary care teams with as many features of the recommended PACT as possible to ensure access to high-quality care, no matter the conditions under which a clinic may be operating at any given time. Future work should examine the impact of staffing levels by specific job role to further optimize primary care team staffing configurations and prepare for future public health emergencies like COVID-19.

## Data Availability

Per VA data security policies, the data must be stored either in secure, shared workspaces or on VA-owned servers not accessible to the public. A limited, deidentified data set, available upon written request made to the PI of the study, will be made available.

